# Immunogenicity and safety of a monovalent Omicron XBB.1.5 SARS-CoV-2 recombinant spike protein vaccine in previously unvaccinated, SARS-CoV-2 seropositive participants: primary day-28 analysis of a phase 2/3 open-label study

**DOI:** 10.1101/2024.10.25.24316129

**Authors:** Katia Alves, Alex Kouassi, Joyce S. Plested, Raj Kalkeri, Katherine Smith, Muneer Kaba, Joy Nelson, Mingzhu Zhu, Shane Cloney-Clark, Zhaohui Cai, Raburn M. Mallory, Fernando Noriega, the 2019nCoV-313 Study

## Abstract

**Background:** Most of the population has been infected with SARS-CoV-2 and, thus, is primed by natural exposure. As such, it was assessed whether a single dose of the monovalent XBB.1.5 vaccine, NVX-CoV2601, elicited a comparable immune response to XBB.1.5 in seropositive unvaccinated participants to that in previously vaccinated participants, thereby allowing the former to forego a two-dose primary series.

**Methods:** In this phase 2/3, open-label, single-arm study (2019nCoV-313/NCT05975060 [part 2]), vaccine-naive participants ≥18 years with previous SARS-CoV-2 infection received one dose of NVX-CoV2601. This analysis compared the 28-day immunogenicity and safety of NVX-CoV2601 in vaccine-naive and previously vaccinated (≥3 prior mRNA-based vaccines, from 2019nCoV-313 part 1) participants.

Noninferiority of neutralizing antibody (nAb) response in vaccine-naive versus vaccinated participants was the primary objective. The day-28 geometric mean titer (GMT) ratio (GMTR) and seroresponse rate (SRR; percentage of participants with a ≥4-fold rise in antibody response from baseline) were measured, and safety was assessed.

**Results:** Of the participants enrolled from September 11–November 15, 2023, per-protocol sets included 306/338 (90.5%) vaccine-naive and 309/332 (93.1%) vaccinated participants. At day 28, adjusted GMTs (95% CI) against XBB.1.5 in the vaccine-naive and vaccinated groups were 1491.5 (1277.5–1741.4) and 841.4 (723.9–978.0), respectively. The vaccine-naïve—vaccinated nAb GMTR was 1.8 (95% CI 1.43–2.20) and SRRs were 74.3% and 64.3% for vaccine-naive and vaccinated participants, respectively (SRR difference: 10.0 [95% CI 2.6–17.4]). No new safety signals or events of special interest were reported.

**Conclusions:** A single dose of NVX-CoV2601 in vaccine-naive participants with a history of SARS-CoV-2 infection elicited a robust neutralizing antibody response that was noninferior to that observed in vaccinated participants. The vaccine was well-tolerated. These data support the use of NVX-CoV2601 as a single dose, regardless of prior vaccination history.

**Trial registration:** NCT05975060

## 1. Introduction

Variants of severe acute respiratory syndrome coronavirus (SARS-CoV-2) have evolved rapidly.^1,2^ The high protective efficacy (≥90%) provided by prototype COVID-19 vaccines (BNT162b2 [Pfizer-BioNTech],^3^ mRNA-1273 [Moderna],^4^ and Matrix-M™–adjuvanted, recombinant spike (rS) protein vaccine NVX-CoV2373 [Nuvaxovid™; Novavax, Inc.]^5,6^) was demonstrated against ancestral SARS-CoV-2 or early variants. Notably, immunity elicited by vaccination or natural infection wanes, and the emergence of immune-evasive variants^7^ suggests the need for updates to vaccine strain composition. The World Health Organization (WHO), European Medicines Agency, and United States Food and Drug Administration (US FDA) recommended updating COVID-19 vaccines to have a monovalent XBB.1.5 composition for the 2023–2024 season.^8–10^ In consideration of this guidance, a monovalent XBB.1.5 subvariant–specific vaccine (NVX-CoV2601) was produced based on the same rS protein technology used for the authorised prototype, NVX-CoV2373. NVX-CoV2601 received authorisation for use in Canada, the European Union (EU), the US, and by the WHO in 2023.^11–14^

Immunogenicity and safety of NVX-CoV2601 are being evaluated in the phase 2/3, open-label 2019nCoV-313 study (NCT05975060) in two populations representative of the US population vaccination status (part 1: previously vaccinated adults who received ≥3 doses of an mRNA vaccine; part 2: vaccine- naive adults who previously had a SARS-CoV-2 infection, as determined by the presence of anti-nucleocapsid [anti-N] antibodies). Results from part 1 showed that in previously vaccinated participants, NVX-CoV2601 induced superior antibody responses to XBB.1.5 compared with those observed in a historical control group of participants who received the prototype vaccine, NVX-CoV2373.^15^

While an estimated 70% of the US population has completed a primary COVID-19 vaccination series as of December 31, 2023, approximately 20% had not received any COVID-19 vaccine doses.^16^ Considering this population, part 2 of the 2019nCoV-313 study aimed to evaluate safety and immunogenicity of a single dose of NVX-CoV2601 administered to unvaccinated participants with a prior SARS-CoV-2 infection. In this case, the unvaccinated population should have some baseline immunity from previous infection, as infection-induced seroprevalence is widespread.^17,18^ A nationwide survey conducted from October–December 2023 showed that ∼87% of the US population (age ≥16 years) had infection-induced seroprevalence (i.e., anti-SARS-CoV-2 spike or nucleocapsid antibodies).^17^ In this setting, it is possible that a single dose of the updated Matrix-M™–adjuvanted, rS protein vaccine would result in adequate and robust immunogenicity in a population whose immune system was not primed by prior COVID-19 vaccination.^18,19^

Here, we report results from the primary day-28 analysis from part 2 of the 2019nCoV-313 study.

In this analysis, neutralizing antibody (nAb) responses elicited by a single dose of NVX-CoV2601, administered to unvaccinated participants with evidence of prior SARS-CoV-2 infection, were evaluated for noninferiority to observed responses to a single dose of NVX-CoV2601 in vaccinated participants who had previously received ≥3 doses of a COVID-19 mRNA vaccine.

## 2. Methods

### 2.1 Study design and participants

Part 2 of the phase 2/3, open-label, single-arm 2019nCoV-313 study (NCT05975060) is being conducted across 30 US sites and has enrolled medically stable males and non-pregnant females aged ≥18 years who were COVID-19 vaccine–naive and had a clinical history of COVID-19–like disease in the previous year. Prior COVID-19–like disease occurrence was collected per participant disclosure at enrollment; time since illness details were not provided. Participants with baseline SARS-CoV-2 seropositivity (anti-N serum IgG) were assessed for the primary immunogenicity endpoint (**Figure S1**). Exclusion criteria included receipt of any investigational vaccine within 90 days before study vaccination and/or receipt of influenza vaccination ≤14 days or of any other approved vaccination ≤30 days before the study vaccination; history of myocarditis/pericarditis; anaphylaxis to any vaccine or allergy to vaccine components; alcohol abuse within 2 years of study vaccination; autoimmune or immunodeficiency disease/condition requiring ongoing therapy; or active cancer treated within 3 years of the first study vaccination. This planned primary analysis includes data through day 28. Additional study details can be found in the study protocol in Appendix B.

The comparator group came from part 1 of the 2019nCoV-313 study, conducted separately, and consisted of participants who had previously received ≥3 doses of mRNA-1273 or BNT162b2 monovalent or bivalent vaccines, with the last dose administered ≥90 days before study vaccination^15^, hereafter referred to as the vaccinated group. Aside from criteria related to previous COVID-19 vaccination and SARS-CoV-2 seropositive status, parts 1 and 2 of the study had the same eligibility criteria.

The study protocol was approved by Advarra, Inc. (Columbia, MD, USA), and the study was conducted according to the principles of the International Conference on Harmonization Good Clinical Practice Guideline, adopting the principles of the Declaration of Helsinki, as well as all applicable national, state, and local laws and regulations. All participants provided written informed consent.

### 2.2 Procedures

Participants in both groups received a single dose of NVX-CoV2601 on day 0 (**Figure S1**) and remained on study for data collection, with scheduled visits at days 28, 90 (phone visit), and 180.

NVX-CoV2601 (5 µg SARS-CoV-2 rS protein from XBB.1.5 and 50 µg Matrix-M^™^ adjuvant) was administered via a 0.5-mL intramuscular injection. Study vaccine was manufactured by the Serum Institute of India, Pvt., Ltd. (Pune, India) in collaboration with Novavax, Inc. Participants in parts 1 and 2 received doses of NVX-CoV2601 from the same vaccine lot and remained in the clinic or under observation for ≥15 min post vaccination for monitoring.

Safety was evaluated throughout the study. Participants completed an eDiary to record solicited reactogenicity for 7 days post vaccination. Reports of unsolicited treatment-emergent adverse events (TEAEs) were collected for 28 days following vaccination. Serious TEAEs (SAEs), treatment-related medically attended adverse events (MAAEs), and adverse events of special interest (AESIs) were collected through day 180 (end of study).

SARS-CoV-2 polymerase chain reaction (PCR) swab testing was performed on day 0. Baseline serostatus was determined through qualitative detection of SARS-CoV-2 anti-N antibodies identified in blood samples collected on day 0; an additional sample was collected on day 28. Blood samples for central evaluation of immunogenicity (Novavax Clinical Immunology, Gaithersburg, MD, USA) were collected on days 0 and 28, with additional samples to be collected on day 180. Serum was analyzed using a validated pseudovirus neutralization assay with an inhibitory dilution of 50% (ID),^20^ anti-rS immunoglobulin G (IgG) enzyme-linked immunosorbent assay (ELISA),^21^ and anti-N serology.^22^ nAb responses to emerging SARS-CoV-2 strains were assessed with fit-for-purpose pseudovirus neutralization assays in a subset of participant samples that had sufficient serum remaining for testing, and were selected through a random number generator.

### 2.3 Outcomes

The primary objective was to determine whether NVX-CoV2601 elicited a noninferior nAb response in SARS-CoV-2–seropositive vaccine-naive participants compared with vaccinated participants. Co-primary endpoints were day-28 nAb geometric mean titers (GMTs) and the proportion of participants who seroconverted (seroresponse rate [SRR]) based on ID_50_ titers to Omicron XBB.1.5). SRR was defined as the percentage of participants with a ≥4-fold rise in antibody response from baseline. IgG antibody responses to XBB.1.5 were evaluated as a secondary endpoint. Post-hoc analyses included nAb responses by age group and responses to emerging SARS-CoV-2 variants, evaluated in a subset of the per-protocol analysis set.

The secondary objective to evaluate the safety of NVX-CoV2601 used the following endpoints: solicited reactogenicity through 7 days post vaccination; unsolicited TEAEs through day 28; and related MAAEs, AESIs and SAEs through day 180. TEAEs were coded (Medical Dictionary for Regulatory Activities, version 26.0) and graded (FDA toxicity grading scale). Solicited reactogenicity events included local (pain, tenderness, redness, and swelling) and systemic (fever, nausea/vomiting, headache, fatigue/malaise, myalgia, and joint pain) reactions. MAAEs were any TEAE leading to an unscheduled healthcare practitioner visit. AESIs included potential immune-medicated medical conditions, myocarditis, pericarditis, and/or COVID-19–specific complications. SAEs were TEAEs associated with death; hospitalization; persistent or significant incapacity or substantial disruption to normal life; congenital anomaly; or other serious, important medical events.

### 2.4 Statistical analysis

Noninferiority of NVX-CoV2601 in vaccine-naive versus vaccinated participants was based on FDA industry guidance^23^ and required meeting two hypotheses: (1) noninferiority based on a GMT ratio (GMTR) with a lower bound of the two-sided 95% confidence interval (CI) exceeding 0.67, and (2) noninferiority based on an SRR difference with the lower bound of the two-sided 95% CI greater than – 10%. The 28-day immunogenicity assessments were completed in the per-protocol analysis set, which included all participants who received the study vaccine and had no major protocol violations or events considered clinically relevant to the immune response (e.g., PCR-positive on day 0) before the data-extraction date and who had days 0 and 28 serology. Participants in the vaccine-naive group were excluded from the per-protocol analysis set if their baseline anti-N result was negative or missing.

A sample size of 330 participants was determined based on the co-primary endpoints.

Assumptions to achieve at least 95% power for GMTR noninferiority were: a standard deviation of 0.6 for log-transformed neutralization titers, a 25% attrition rate, and an overall one-sided Type I error rate of 2.5%. The sample size of 330 participants provided a likelihood of detecting rare TEAEs, with probability– risk combinations determined from the actual sample size (not the sample size adjusted for the attrition rate). Specifically, there was a probability of detecting at least one TEAE of 91.89% if the true incidence was 0.010; probabilities were 89.57%, 86.57%, 82.73%, 77.79%, and 71.44% if the true incidences were 0.009, 0.008, 0.007, 0.006, and 0.005, respectively. Secondary immunogenicity endpoints were descriptively summarised.

GMTs and geometric mean ELISA units (GMEUs) were calculated as the antilog of the mean of the log-transformed titers at day 28 to generate a normal distribution. An analysis of covariance with vaccine group (vaccine-naive versus vaccinated) as a fixed effect and baseline value (day 0) as covariate was performed to estimate the adjusted GMT and GMTR, and account for baseline differences among the groups. The same methodology was used to estimate adjusted GMEUs and GMEU ratios (GMEURs). The mean difference and corresponding CI limits between vaccine groups were exponentiated to GMTRs and GMEURs and corresponding 95% CIs. The 95% CIs for adjusted GMTs and GMTRs were calculated based on the *t*-distribution of the log-transformed values, then back transformed to the original scale.

SRR two-sided exact binomial 95% CIs were calculated using the Clopper-Pearson method; SRR differences (vaccine-naive–vaccinated) were computed along with their corresponding two-sided 95% CIs, based on the method of Miettinen and Nurminen.

The safety analysis set included all participants who provided consent and received study vaccine; the safety set was analysed as actually treated. The descriptive safety analysis included summaries of the number and percentage of participants with solicited (local and systemic) TEAEs through 7 days and unsolicited AEs through 28 days after the study vaccination. Two-sided exact 95% CIs using the Clopper–Pearson method were included. The interim analysis included a descriptive summary of all MAAEs, AESIs, and SAEs through 28 days post study vaccination. SAS® software (version 9.4; SAS Institute Inc., Cary, NC, USA) was used for all statistical analyses.

## 3. Results

### 3.1 Participants

Between September 11, 2023 and November 15, 2023, 338/419 (80.7%) participants were enrolled in part 2 of the study, after eligibility screening; there were 81 screen failures (**Figure 1**). At the data extraction date (February 21, 2024), 322/338 (95.3%) participants were in follow-up and 16/338 (4.7%) discontinued (reasons: 4 [1.2%] participant withdrawal; 10 [3.0%] lost to follow-up; 1 [0.3%] investigator decision [0.3%]; 1 [0.3%] other). No participant discontinued the study because of a TEAE. All 338 participants were included in the safety analysis set. The per-protocol analysis set included 306/338 (90.5%) participants; reasons for exclusion were negative baseline anti-N (23/338; 6.8%), positive baseline PCR (5/338; 1.5%), and protocol violation (5/338; 1.5%; one patient had a protocol violation and positive PCR result). For the vaccinated group, 329/332 (99.1%) participants were in follow-up at the data extraction date. All 332 participants were included in the safety analysis set, and the per-protocol analysis set included 309/332 (93.1%) participants. The vaccinated group (from part 1 of the study) was enrolled from September 7–8, 2023^15^ and included 332 and 309 participants in the safety and per-protocol analysis sets, respectively.

**Figure 1.**
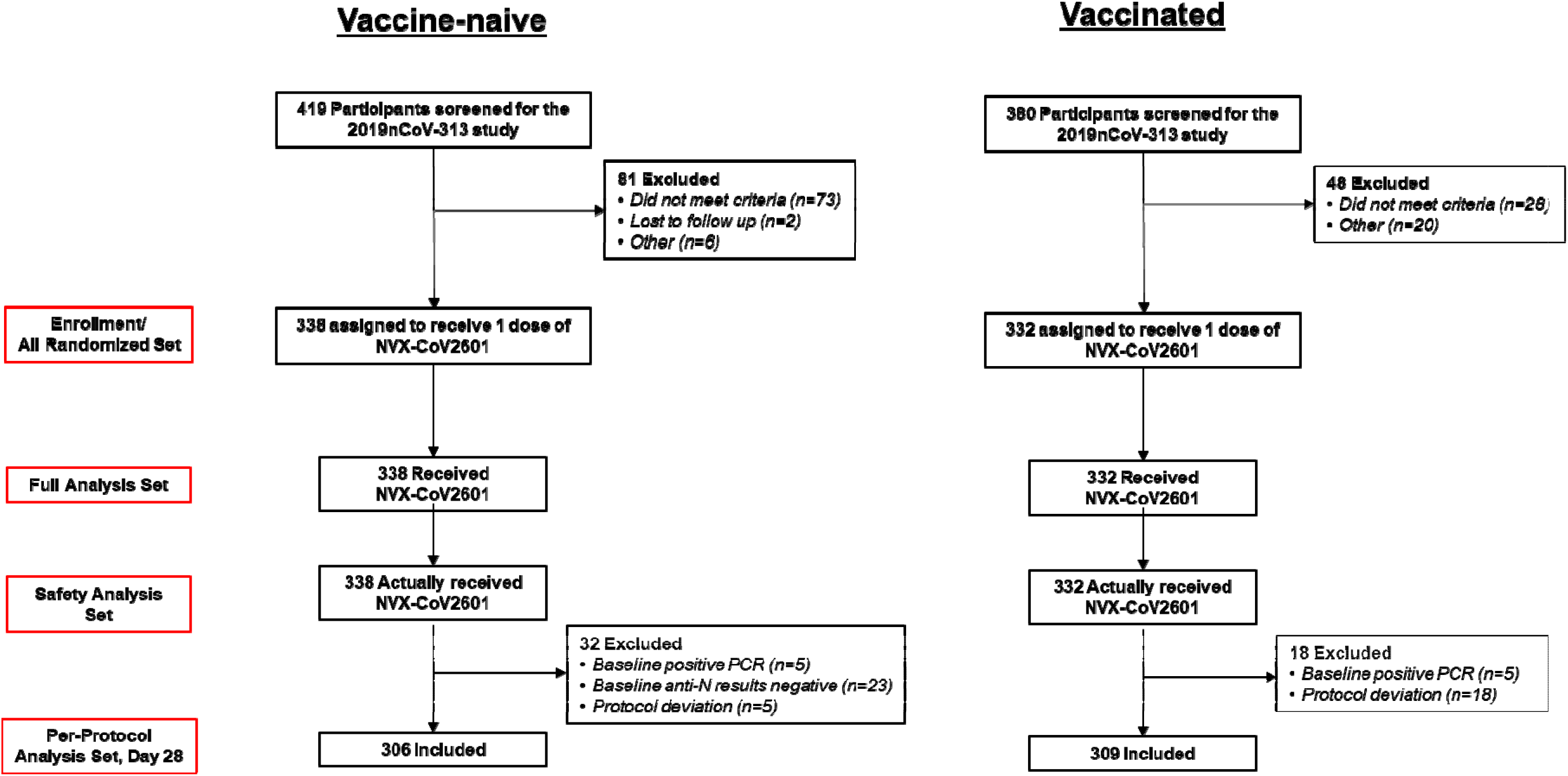
CONSORT diagram. Participants in the previously vaccinated (comparator) group were enrolled in part 1 of the 2019nCoV- 313 study. Participants may have been excluded for more than one reason.

Some differences were observed in participant demographics for the vaccine-naive and vaccinated groups (**Table1**). In the per-protocol analysis set, median age (IQR) was 38 years (31–49) in the vaccine-naive group and 53 years (40–65) in the vaccinated group. Most participants in the vaccine- naive group identified as either White (152/306 [49.7%]) or Black/African American (132/306 [43.1%]). In the vaccinated group, most participants identified as White (231/309 [74.8%]), and a smaller proportion identified as Black/African American (47/309 [15.2%]). Most participants in the vaccinated group had previously received three (129/309 [41.7%]) or four (109/309 [35.3%]) doses of an mRNA-based COVID-19 vaccine. Baseline demographics and participant characteristics were similar between the per-protocol and safety analysis sets (**Table S1**).

**Table 1.** Participant baseline demographics and characteristics in the per-protocol analysis set.

|  | **Vaccine-naïve**<br>**(N=306)** | **Vaccinated**<br>**(N=309)** |
| --- | --- | --- |
| **Age, years** |  |  |
| Mean (SD) | 40 (12.91) | 52.1 (16.10) |
| Median (IQR) | 38 (31–49) | 53 (40–65) |
| **Age, category, n (%)** |  |  |
| 18 to 54 years | 263 (85.9) | 163 (52.8) |
| ≥55 years | 43 (14.1) | 146 (47.2) |
| **Sex, n (%)** |  |  |
| Female | 177 (57.8) | 192 (62.1) |
| Male | 129 (42.2) | 117 (37.9) |
| **Race, n (%)** |  |  |
| White | 152 (49.7) | 231 (74.8) |
| Black/African American | 132 (43.1) | 47 (15.2) |
| Native American/Alaska Native | 6 (2.0) | 6 (1.9) |
| Asian | 2 (0.7) | 12 (3.9) |
| Native Hawaiian/other Pacific Islander | 1 (0.3) | 2 (0.6) |
| Multiple | 5 (1.6) | 3 (1.0) |
| Other | 2 (0.7) | 1 (0.3) |
| Unknown/not reported | 6 (2.0) | 7 (2.2) |
| **Ethnicity, n (%)** |  |  |
| Not Hispanic/Latino | 224 (73.2) | 245 (79.3) |
| Hispanic/Latino | 80 (26.1) | 61 (19.7) |
| Not reported | 2 (0.7) | 3 (1.0) |
| **Previous COVID-19, n (%)** | 302 (98.7) | 5 (1.6) |
| **PCR, n (%)** |  |  |
| Negative | 306 (100)* | 309 (100)<sup>†</sup> |
| Positive | 0 | 0 |
| **Anti-N/PCR status, n (%)<sup>‡</sup>** |  |  |
| Positive | 306 (100) | 215 (69.6) |
| Negative | 0 | 94 (30.4) |
| **Time between previous COVID-19 vaccine and study vaccination, days** |  |  |
| Mean (SD) | NA | 438.2 (177.32) |
| Median (IQR) | NA | 361 (314–623) |

\*Five participants with missing PCR status at baseline were imputed as negative in part 2.
<sup>†</sup>Two participants with missing PCR status at baseline were imputed as negative in part 1.
<sup>‡</sup>Participants with a positive result for either anti-N or PCR are reported.

### 3.2 Immunogenicity

The study met the two co-primary objectives; NVX-CoV2601 elicited a noninferior nAb response in vaccine-naive participants compared with vaccinated participants (**Table2**). At day 0, nAb GMTs against XBB.1.5 were lower in the vaccine-naive group (67.6 [95% CI 56.8–80.4]) compared with the vaccinated group (120.8 [101.5–143.8]) (**Figure 2A**). At day 28, the increase in nAb GMTs from day 0 was 19.3-fold in the vaccine-naive group (GMT: 1303.7 [1087.4–1563.0]; adjusted GMT: 1491.5 [1277.5–1741.4]) and 7.9-fold in the vaccinated group (GMT: 955.5 [95% CI 814.0–1121.4]; adjusted GMT: 841.4 [723.9–978.0]). The nAb response was similar between groups (**Figure 2B**). The GMTR between the groups at day 28 was 1.8 (95% CI 1.43–2.20) (**Figure 2C**). From day 0 to 28, SRRs were 74.3% and 64.3% for the vaccine-naive and vaccinated groups, respectively, with an SRR difference of 10.0 (95% CI 2.6– 17.4). When assessed by age group, nAb GMTs (95% CI) in participants aged 18–54 years increased from day 0 to 28 by 18.4-fold in the vaccine-naive group (GMT: 1208.3 [993.1–1470.2]; adjusted GMT: 1337.6 [1125.3–1589.9]) and 8.4-fold in the vaccinated group (GMT: 1027.9 [829.5–1090.9]; adjusted GMT: 880.6 [710.9–1090.9]) (**Figure S2**). Increases in nAb GMTs were also observed in participants aged ≥55 years: 25.6-fold in the vaccine-naive group (GMT: 2009.9 [1250.4–3231.0]; adjusted GMT: 2364.3 [1633.0–3423.1]) and 7.4-fold in the vaccinated group (GMT: 880.5 [95% CI 691.2–1121.5]; adjusted GMT: 838.8 [685.6–1026.1]). The anti-rS IgG GMEU on day 0 in the vaccine-naive group was 5218.1 (95% CI 4339.6–6274.5), whereas it was 23701.1 (21042.3–26695.8) in the vaccinated group, a median of ∼12 months after their most recent COVID-19 vaccination (**Table S2**). On day 28, GMEU levels increased in both groups (adjusted GMEUs: 60691.2 [54585.2–67480.3] and 79761.5 [73603.5–86434.7], respectively). Descriptive results of the anti-rS IgG analyses were consistent with the nAb analyses, with similar anti-rS IgG titers and SRRs between groups.

**Figure 2.**
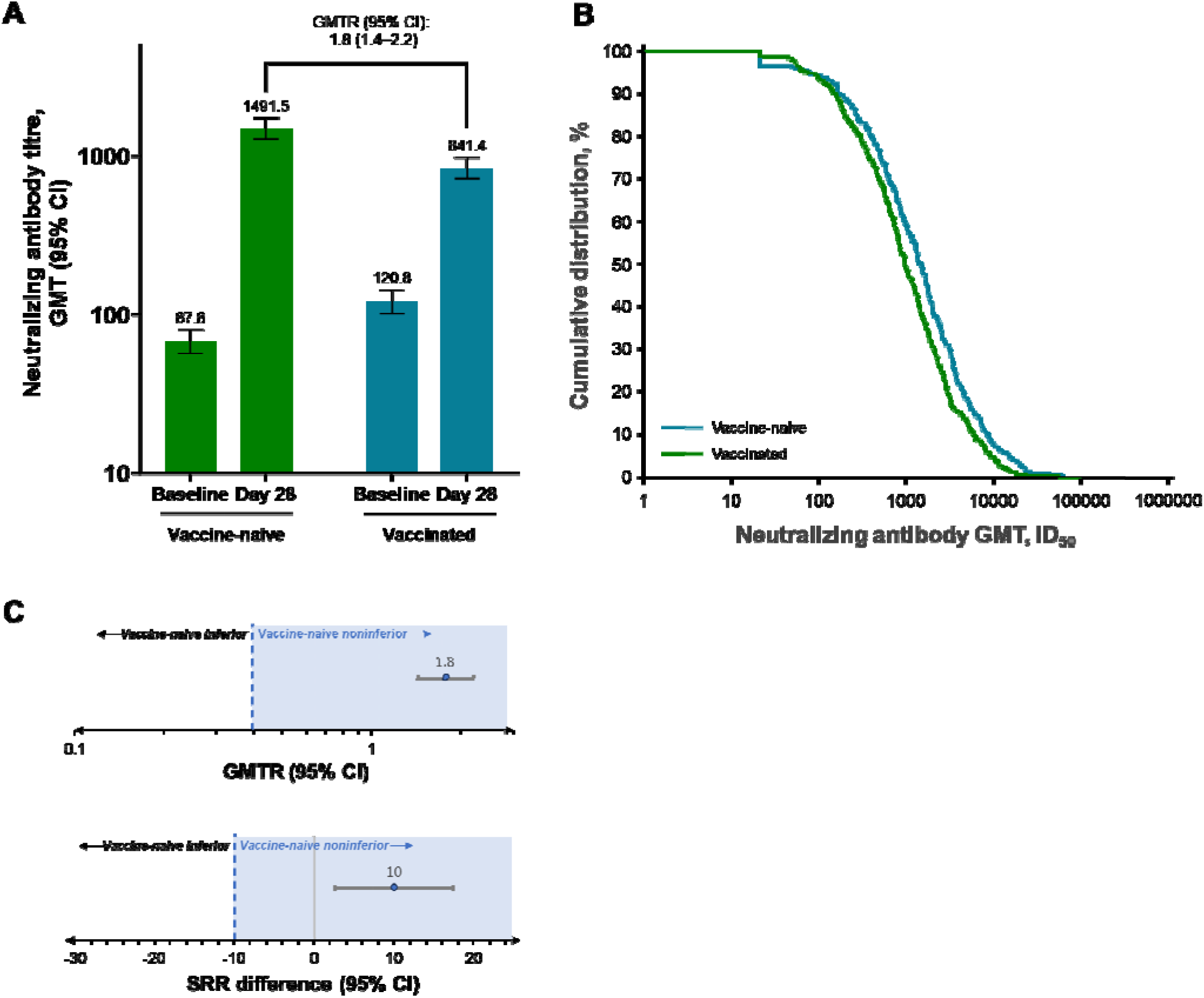
Neutralizing antibody responses to XBB.1.5 at days 0 and 28 post study vaccination (per- protocol analysis set) (A) GMTs of nAb responses to XBB.1.5 in vaccine-naive and previously vaccinated participants. GMT values are shown on a log scale *y*-axis, and corresponding adjusted GMT values are shown above each respective bar. GMTRs (95% CI) are included comparing bracketed bars (noninferiority criterion: lower bound of the two-sided 95% CI for the GMTR > 0.67). (B) Cumulative distribution of nAb ID_50_ GMTs. (C) Top panel: GMTR (log-scale GMT ratio of vaccine-naive to previously vaccinated) with a lower bound of the two-sided 95% CI greater than 0.67 indicating noninferiority of vaccine-naive vs previously vaccinated. Bottom panel: a SRR difference (vaccine-naive – previously vaccinated) with a lower bound of the two-sided 95% CI greater than –10% indicating vaccine-naive non-inferiority to previously vaccinated. The dashed vertical lines represent the noninferiority bounds, and the blue box indicates the noninferiority region for vaccine-naive.

**Table 2.**
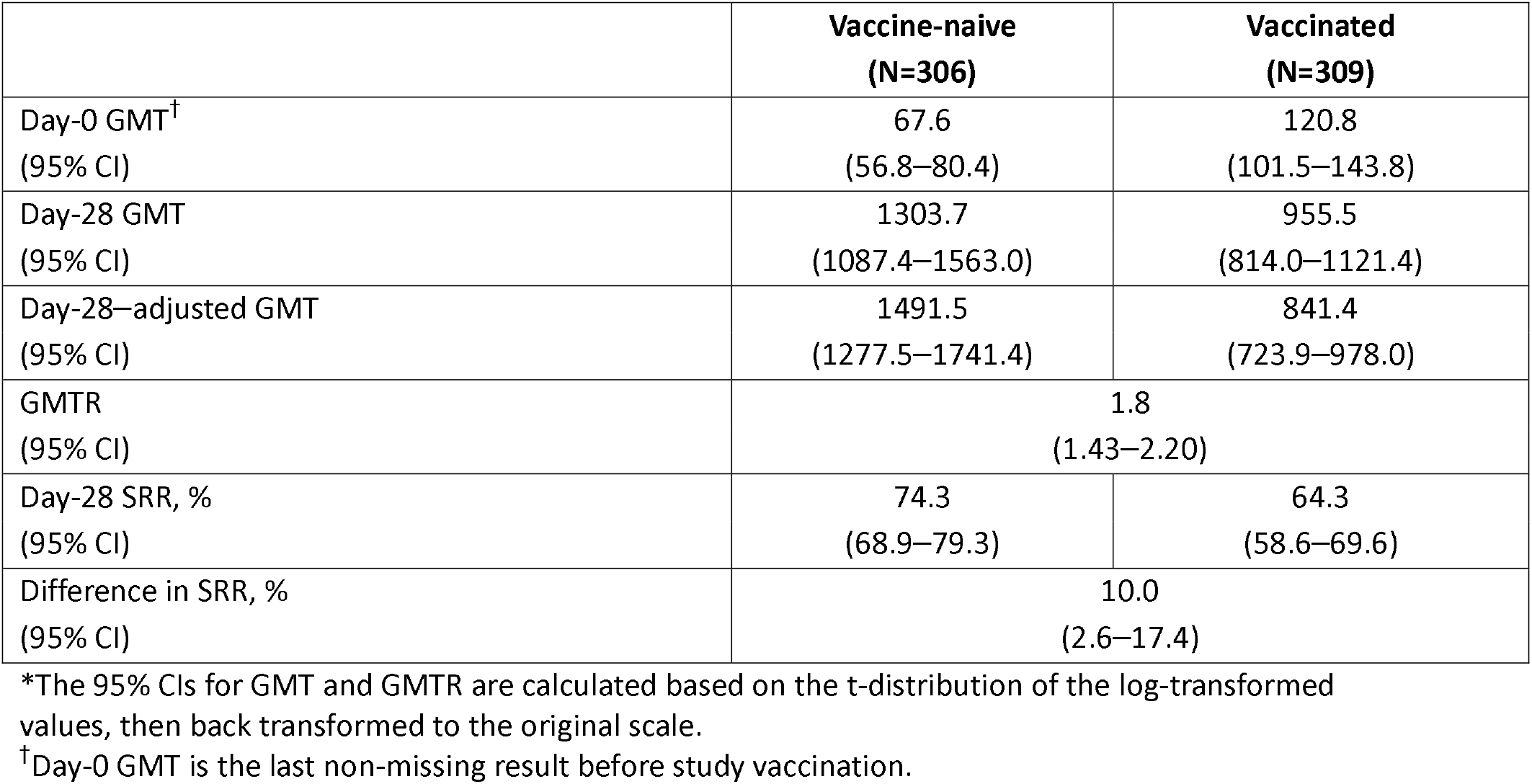
Neutralizing antibody titers (GMT* ID_50_) for Omicron XBB.1.5 (Primary endpoint; per-protocol analysis set)

|  | **Vaccine-naïve**<br>**(N=306)** | **Vaccinated**<br>**(N=309)** |
| --- | --- | --- |
| Day-0 GMT<sup>†</sup><br>(95% CI) | 67.6<br>(56.8–80.4) | 120.8<br>(101.5–143.8) |
| Day-28 GMT<br>(95% CI) | 1303.7<br>(1087.4–1563.0) | 955.5<br>(814.0–1121.4) |
| Day-28—adjusted GMT<br>(95% CI) | 1491.5<br>(1277.5–1741.4) | 841.4<br>(723.9–978.0) |
| GMTR<br>(95% CI) | 1.8<br>(1.43–2.20) |  |
| Day-28 SRR, %<br>(95% CI) | 74.3<br>(68.9–79.3) | 64.3<br>(58.6–69.6) |
| Difference in SRR, %<br>(95% CI) | 10.0<br>(2.6–17.4) |  |

\*The 95% CIs for GMT and GMTR are calculated based on the t-distribution of the log-transformed values, then back transformed to the original scale.
<sup>†</sup>Day-0 GMT is the last non-missing result before study vaccination.

Based on a subset of samples from the vaccine-naive group (n=42), GMTs against additional SARS-CoV-2 variants increased from day 0 to 28 (**Figure 3**). At day 0, the GMT (95% CI) against JN.1 was 51.2 (33.5–78.3), which increased to 486.4 (310.0–763.2) at day 28 (GMFR: 9.5). Day 0 and day 28 GMTs (95% CI), respectively, against KP.2 were 44.2 (28.5–68.6) and 360.4 (232.7–558.2), with a GMFR of 9.1. Among the variants analyzed in this subset, the highest baseline and day 28 titers (and GMFR) were observed against XBB.1.5.

**Figure 3.**
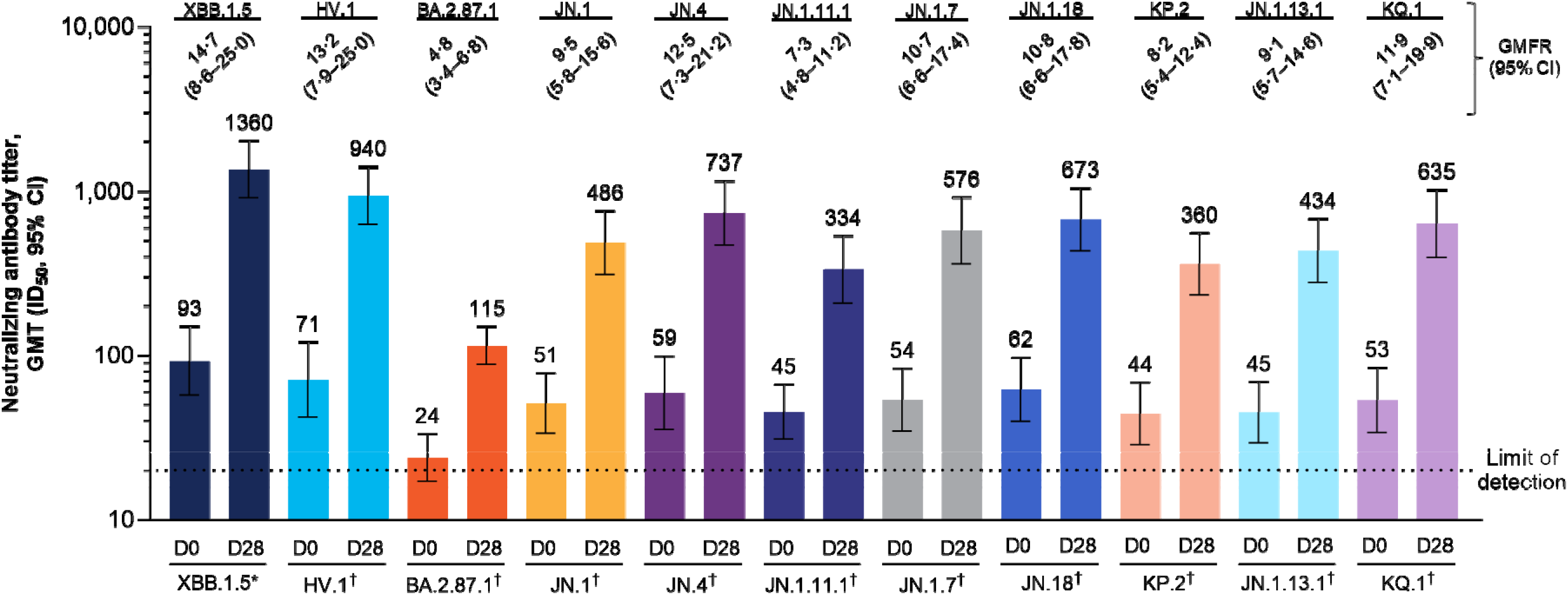
Pseudovirus neutralizing antibody responses to XBB.1.5 and emerging variants at days 0 and 28 post study vaccination in a subset of the vaccine-naive group (Per-protocol analysis subset) Neutralizing antibody responses are shown in a subset of participants from the per-protocol analysis set (n=42). GMT values are shown on a log scale *y*-axis, and corresponding adjusted GMT values are shown above each respective bar. *Validated assays. ^†^Fit for purpose assays.

### 3.3 Safety

Within 7 days of study vaccine administration, 140/338 (41.4%) vaccine-naive participants and 189/332 (56.9%) vaccinated participants reported a local solicited TEAE (**Figure 4**; **Table 3**). The most common (occurring in >20% of participants) solicited local event was pain/tenderness (combined proportion), reported in 140/338 (41.4%) and 186/332 (56.0%) of the vaccine-naive and vaccinated groups, respectively (median duration: 2 days in both groups). Three (0.9%) participants in the vaccine- naive group (grade 3 pain/tenderness) and one (0.3%) in the vaccinated group (grade 3 pain/tenderness) had solicited local TEAEs of grade ≥3.

**Figure 4.**
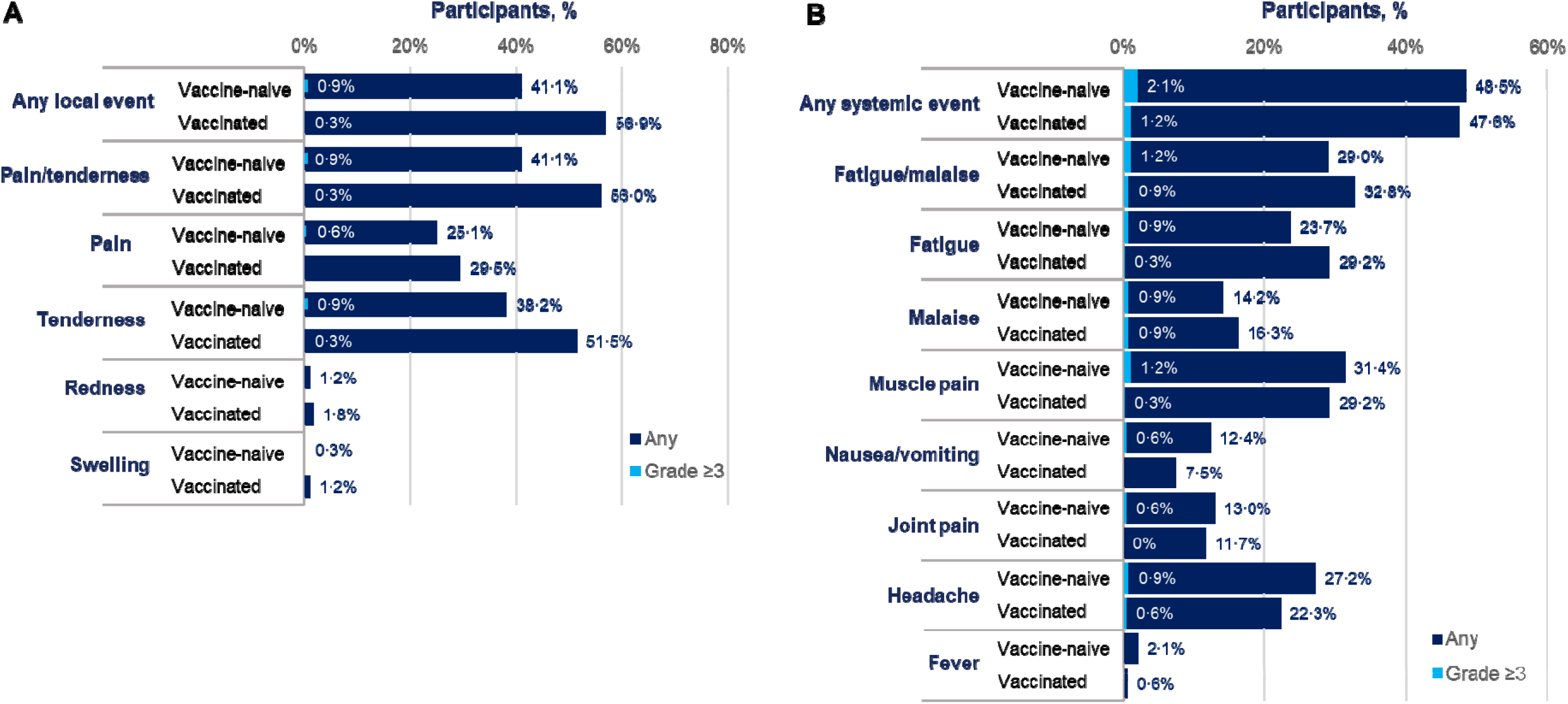
Solicited local and systemic reactogenicity events in all participants (Safety analysis set) Solicited (A) local and (B) systemic TEAEs within 7 days of study vaccination in participants who received study vaccination and completed at least one eDiary entry. The proportion of participants with a grade ≥3 event is shown in gray text within the blue bars.

**Table 3.** Summary of solicited and unsolicited TEAEs in vaccine-naive and previously vaccinated participants (Safety analysis set)

|  | **Vaccine-naïve**<br>**(N=338)** | **Vaccinated**<br>**(N=332)** |
| --- | --- | --- |
| Solicited AEs, n (%) | 202 (59.8) | 227 (68.4) |
| Grade ≥3 | 9 (2.7) | 5 (1.5) |
| Local | 140 (41.4) | 189 (56.9) |
| Grade ≥3 | 3 (0.9) | 1 (0.3) |
| Systemic | 164 (48.5) | 158 (47.6) |
| Grade ≥3 | 7 (2.1) | 4 (1.2) |
| Unsolicited TEAEs, n (%) |  |  |
| Any | 18 (5.3) | 29 (8.7) |
| Treatment-related | 1 (0.3) | 5 (1.5) |
| Severe TEAEs, n (%) | 1 (0.3)* | 2 (0.6)<sup>†</sup> |
| Treatment-related | 0 | 0 |
| Serious TEAE, n (%) | 1 (0.3)* | 2 (0.6)<sup>†</sup> |
| Treatment-related | 0 | 0 |
| Medically attended AE, n (%) | 8 (2.4) | 14 (4.2) |
| Severe | 0 | 2 (0.6)<sup>†</sup> |
| Treatment-related | 0 | 1 (0.3)<sup>†</sup> |
| AESIs, n (%)<sup>‡</sup> | 0 | 0 |
| TEAEs leading to study discontinuation, n (%) | 0 | 0 |
| Death, n (%) | 0 | 0 |

\*The severe/serious TEAE was obstructive pancreatitis.
<sup>†</sup>Severe/serious TEAEs and medically-attended, severe AEs were an appendiceal abscess and a gastrointestinal stromal tumor. The related medically attended AE was hypertension.
<sup>‡</sup>AESIs include potential immune-mediated medical conditions and AEs specific to COVID-19 (i.e., myocarditis/pericarditis).

Solicited systemic TEAEs within 7 days of study vaccine were reported in 164/338 (48.5%) vaccine-naive and 158/332 (47.6%) vaccinated participants. The most common solicited systemic events were fatigue/malaise (vaccine-naive: 98/338 [29.0%]; vaccinated: 109/332 [32.8%]), muscle pain (106/338 [31.4%]; 97/332 [29.3%]), and headache (92/338 [27.2]; 74/332 [22.3%]). Median durations of fatigue/malaise were 2 days in both groups; muscle pain and headache both had median durations of 2 days in the vaccine-naive group and 1 day in the vaccinated group. Few participants reported a solicited event of fever (vaccine-naive: 3/338 [0.9%]; vaccinated: 2/332 [0.6%]). Grade ≥3 solicited systemic TEAEs were reported in 7/338 (2.1%) and 4/332 (1.2%) participants in the vaccine-naive and vaccinated groups, respectively, none of which were grade 4.

Through 28 days post-vaccination, unsolicited TEAEs were reported in 18/338 (5.3%) and 29/332 (8.7%) participants in the vaccine-naive and vaccinated groups, respectively (**Table 3**; **Table S3**). Few participants reported treatment-related unsolicited TEAEs. Among 8/338 (2.4%) and 14/332 (4.2%) participants with an MAAE in the vaccine-naive and vaccinated groups, respectively, one MAAE of grade 2 hypertension in a participant from the vaccinated group was considered treatment-related. SAEs and severe TEAEs were reported in <1% of participants in either group; none were considered treatment-related. There were no reported events of myocarditis, pericarditis, or any other AESI. No participant discontinued the study because of a TEAE, and no deaths were reported.

## 4. Discussion

In this primary day-28 analysis of part 2 of the 2019nCoV-313 study, immunogenicity of a single dose of the monovalent XBB.1.5 vaccine (NVX-CoV2601), resulted in robust immunogenicity in vaccine- naive participants seropositive for SARS-CoV-2 N-protein, and no new safety signals were identified. The co-primary endpoints of noninferiority of the GMTR and SRR difference between vaccine-naive and vaccinated participants, who both received single study doses of NVX-CoV2601, were met.

Robust increases in nAbs were observed for both study groups after receipt of NVX-CoV2601. Notably, a greater increase in titer was observed from day 0 to 28 in the vaccine-naive group (19.3-fold increase in GMT) compared with the vaccinated group (7.9-fold increase). This observation may be related to lower baseline titers and lower median age in the vaccine-naive group, as increasing age has been associated with immunosenescence and decreased vaccine-related immunity.^24^ However, a post-hoc analysis evaluating nAb response by age found that 28-day nAb titers were higher in vaccine-naive compared with previously vaccinated participants in both age groups (18–54 and ≥55 years). To help address reduction in immune titers over time in older adults, an additional dose of the monovalent XBB.1.5 vaccine was recommended by regulatory authorities for the spring of 2024 for older adults and those who are immunocompromised.^25^

The overwhelming majority of the US population has been infected with SARS-CoV-2, received a COVID-19 vaccination, or both.^17,26^ In a nationwide study of US blood donors, 87% had anti-N antibodies indicating a past infection, and combined seroprevalence from infection or vaccination reached 98% by the end of 2023.^17^ In this epidemiologic environment, a single dose of NVX-CoV2601 led to a similar nAb response between vaccine-naive and previously vaccinated participants, suggesting that a single dose was sufficient to augment immunogenicity derived from natural infection. Other studies have shown that one vaccine dose in unvaccinated people post SARS-CoV-2 infection can induce a substantial immune response that is comparable to, if not greater than, that observed in those who have received two vaccine doses.^18,19^ Overall, amidst high population-wide seroprevalence, priming via vaccination (e.g., two doses administered 21 days apart) no longer seems necessary for adequate immunogenicity, and these data from the 2019nCov-313 study support the use of NVX-CoV2601 as a single dose in unvaccinated individuals.

The rapid evolution of SARS-CoV-2 variants led to an updated monovalent XBB.1.5 vaccine composition for 2023–2024^8–15^ and recent authorization of another updated vaccine targeting the JN.1 lineage for 2024–2025^8,27^. Vaccination with a single dose of NVX-CoV2601 elicited robust immunogenicity against XBB.1.5 in vaccine-naive and vaccinated individuals, as well as cross-reactivity with variants that have emerged since XBB.1.5 in the vaccine-naive group (i.e., approximately 10-fold increases in nAb GMTs against JN.1 and lineage subvariants, including KP.2). Cross-reactivity has also been reported separately in the vaccinated group.^15^ These findings supported recommendations for the spring 2024 vaccination campaign in vulnerable individuals such as older adults and those with weakened immune systems.^25^ However, titers against these variants were approximately 50–75% lower compared with the titers against XBB.1.5, suggesting a benefit to periodic COVID-19 vaccine updates to align with predominantly circulating strains, such as that seen for 2024–2025.^27^

This study is an open-label study without randomization. Participants were sequentially enrolled in study groups in two separate parts, which allowed for an earlier evaluation of the safety and immunogenicity of a single dose of NVX-CoV2601 in part 1 (≥3 mRNA previously vaccinated participants) followed by enrollment of vaccine-naive participants with previous SARS-CoV-2 infection in part 2.

Participants were recruited from the same set of study sites and enrollment was sequential; therefore, both populations were exposed to similar SARS-CoV-2 strains circulating at this time (e.g., XBB-lineage^28^), and any differences among the groups likely reflect differences typically observed among COVID-19– vaccinated and –unvaccinated individuals in the general US population. Notably, enrollment of participants who had remained unvaccinated for the 3 years since vaccines against SARS-CoV-2 first became available, and would consent to study vaccination, was anticipated to be a challenge. As such, a limitation of the study is some differences in participant demographics (e.g., age, race, and sex) and prior COVID-19–like disease occurrence between the study groups. Specifically, the vaccine-naive group had larger proportions of participants with demographics associated with vaccine hesitancy, such as younger age and Black/African American race.^29^

In summary, a single dose of NVX-CoV2601 elicited robust immunogenicity in vaccine-naive participants with prior SARS-CoV-2 infection. In this population, immunogenicity of NVX-CoV2601 was noninferior to that observed in participants who previously received ≥3 mRNA vaccinations. Additionally, the safety and reactogenicity profile of the updated vaccine targeting the XBB.1.5 strain, NVX-CoV2601, was consistent with that of the prototype vaccine, NVX-CoV2373^5^. Amidst high seroprevalence in the US, these data support the use of single doses of the Novavax vaccine to elicit immunity against SARS-CoV-2, regardless of prior vaccination history.

## Supporting information

Appendix A for Noriega et al.

## Data Availability

Study information is available online at https://www.clinicaltrials.gov/study/NCT05975060. Requests submitted to the corresponding author will be considered upon publication of this article.

## Acknowledgments

We thank all the study participants who volunteered for this study. We also thank Sharon Glass, MLIS, of Novavax, Inc., for providing literature research support, and Gordon Chau, MS, of Novavax, Inc., for statistical support. Medical writing, editing, and formatting support, under the direction of the authors, was provided by Meredith Kalish, MD, CMPP and Ebenezer M. Awuah-Yeboah, BS, of Ashfield MedComms (US), an Inizio company, and was funded by Novavax, Inc.

## Appendix A. Supplementary data

Supplementary study details and results.

## Appendix B. Study protocol

2019nCoV-313 study protocol.

## Funding

Novavax, Inc. funded the 2019nCoV-313 study and provided the vaccine. Novavax, Inc. collaborated with the investigators on the protocol, data analysis and interpretation, and preparation of this paper.

## CRediT authorship contribution statement

**Katia Alves** : Validation; Writing – reviewing & editing

**Alex Kouassi**: Conceptualization; Formal analysis; Validation; Writing – reviewing & editing

**Joyce S. Plested**: Formal analysis; Writing – reviewing & editing

**Raj Kalkeri**: Methodology; Formal analysis; Writing – reviewing & editing

**Katherine Smith**: Formal analysis; Writing – reviewing & editing

**Muneer Kaba**: Formal analysis; Writing – reviewing & editing

**Joy Nelson**: Project administration; Writing – reviewing & editing

**Mingzhu Zhu**: Formal analysis; Writing – reviewing & editing

**Shane Cloney-Clark**: Formal analysis; Writing – reviewing & editing

**Zhaohui Cai**: Formal analysis; Writing – reviewing & editing

**Raburn M. Mallory**: Conceptualization; Writing – reviewing & editing

**Fernando Noriega**: Conceptualization; Validation; Writing – reviewing & editing

## Data Statement

Study information is available online at https://www.clinicaltrials.gov/study/NCT05975060. Requests submitted to the corresponding author will be considered upon publication of this article. The study protocol is available in Appendix B.

## Declaration of Competing Interest

Katia Alves reports a relationship with Novavax Inc that includes: employment and equity or stocks. Alex Kouassi reports a relationship with Novavax Inc that includes: employment and equity or stocks. Joyce S. Plested reports a relationship with Novavax Inc that includes: employment and equity or stocks. Raj Kalkeri reports a relationship with Novavax Inc that includes: employment and equity or stocks.

Katherine Smith reports a relationship with Novavax Inc that includes: employment and equity or stocks. Muneer Kaba reports a relationship with Novavax Inc that includes: employment and equity or stocks.

Joy Nelson reports a relationship with Novavax Inc that includes: employment and equity or stocks. Mingzhu Zhu reports a relationship with Novavax Inc that includes: employment and equity or stocks. Shane Cloney-Clark reports a relationship with Novavax Inc that includes: employment and equity or stocks. Zhaohui Cai reports a relationship with Novavax Inc that includes: employment and equity or stocks. Raburn M. Mallory reports a relationship with Novavax Inc that includes: employment and equity or stocks. Fernando Noriega reports a relationship with Novavax Inc that includes: employment and equity or stocks. This study was funded by Novavax, Inc. The authors received medical writing and editorial support funded by the study sponsor. If there are other authors, they declare that they have no known competing financial interests or personal relationships that could have appeared to influence the work reported in this paper.

## Abbreviations

AESI: adverse event of special interest
ELISA: enzyme-linked immunosorbent assay
GMEU: geometric mean ELISA unit
GMEUR: geometric mean ELISA unit ratio
GMT: geometric mean titer
GMTR: geometric mean titer ratio
IQR: interquartile ratio
LB: lower bound
MAAE: medically attended adverse event
N: nucleocapsid
nAb: neutralizing antibody
SAE: serious adverse event
SRR: seroresponse rate
TEAE: treatment-emergent adverse event

## Notes

### Clinical Trial

NCT05975060

