## Appendix A for Noriega et al. for "Immunogenicity and safety of a monovalent Omicron XBB.1.5 SARS-CoV-2 recombinant spike protein vaccine in previously unvaccinated, SARS-CoV-2 seropositive participants: primary day-28 analysis of a phase 2/3 open-label study"

**Table of Contents**

**Figure S1.** Study design	3

**Figure S2.** Disposition	4

**Figure S3.** Neutralising antibody responses to XBB.1.5 at days 0 and 28 post study vaccination by age group (Per-protocol analysis set)	5

**Table S1.** Participant baseline demographics and characteristics in the safety analysis set	6

**Table S2.** Anti-rS IgG responses against XBB.1.5 (Per-protocol analysis set)	7

**Table S3.** Unsolicited TEAEs through 28 days post study vaccination (Safety analysis set)	8

**2019nCoV-313 study investigators**	9

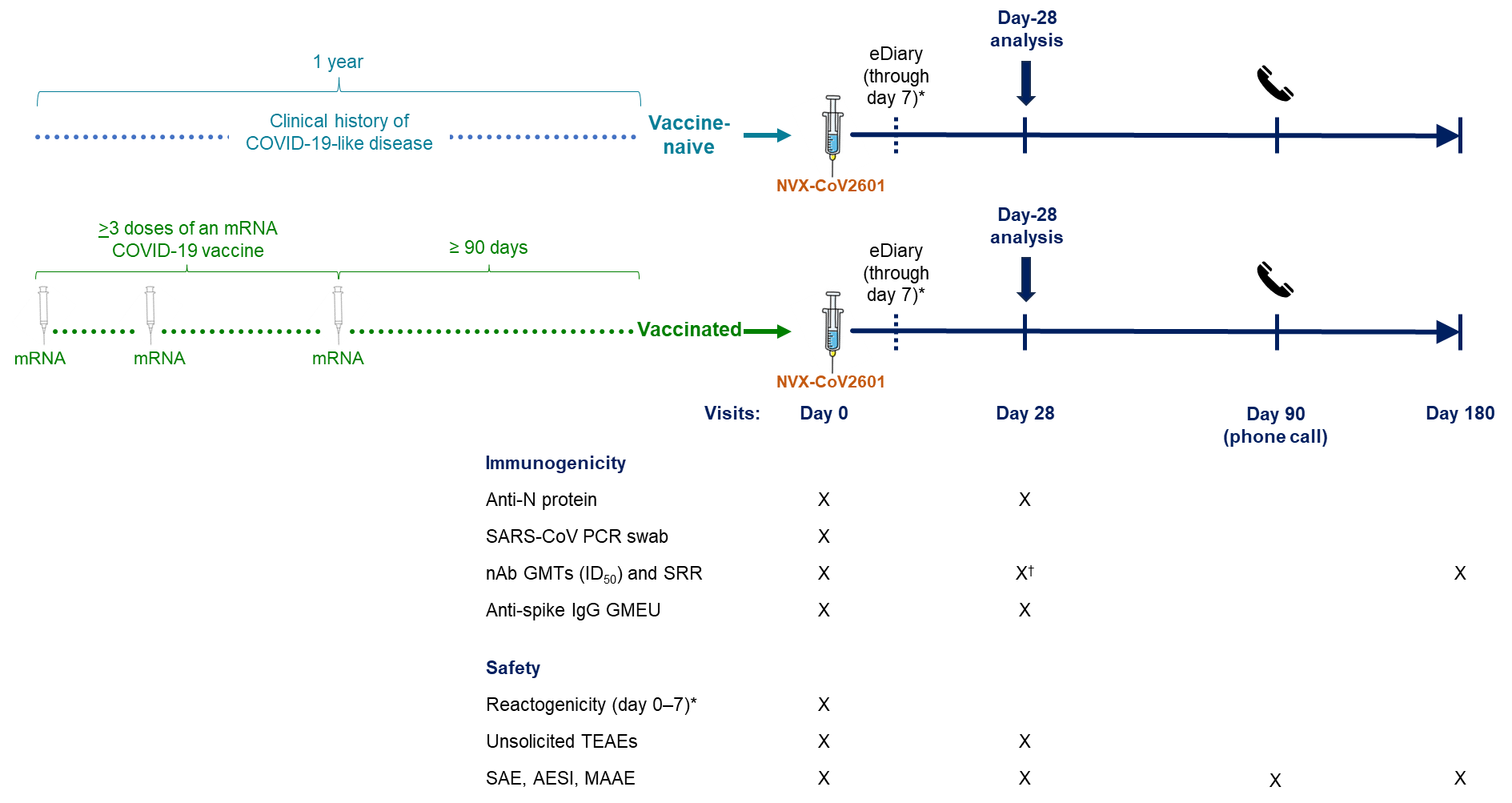

**Figure S1.** Study design

^*^Reactogenicity events consisted of solicited TEAEs occurring ≤7 days post vaccination, collected via eDiary. ^†^Primary endpoint.

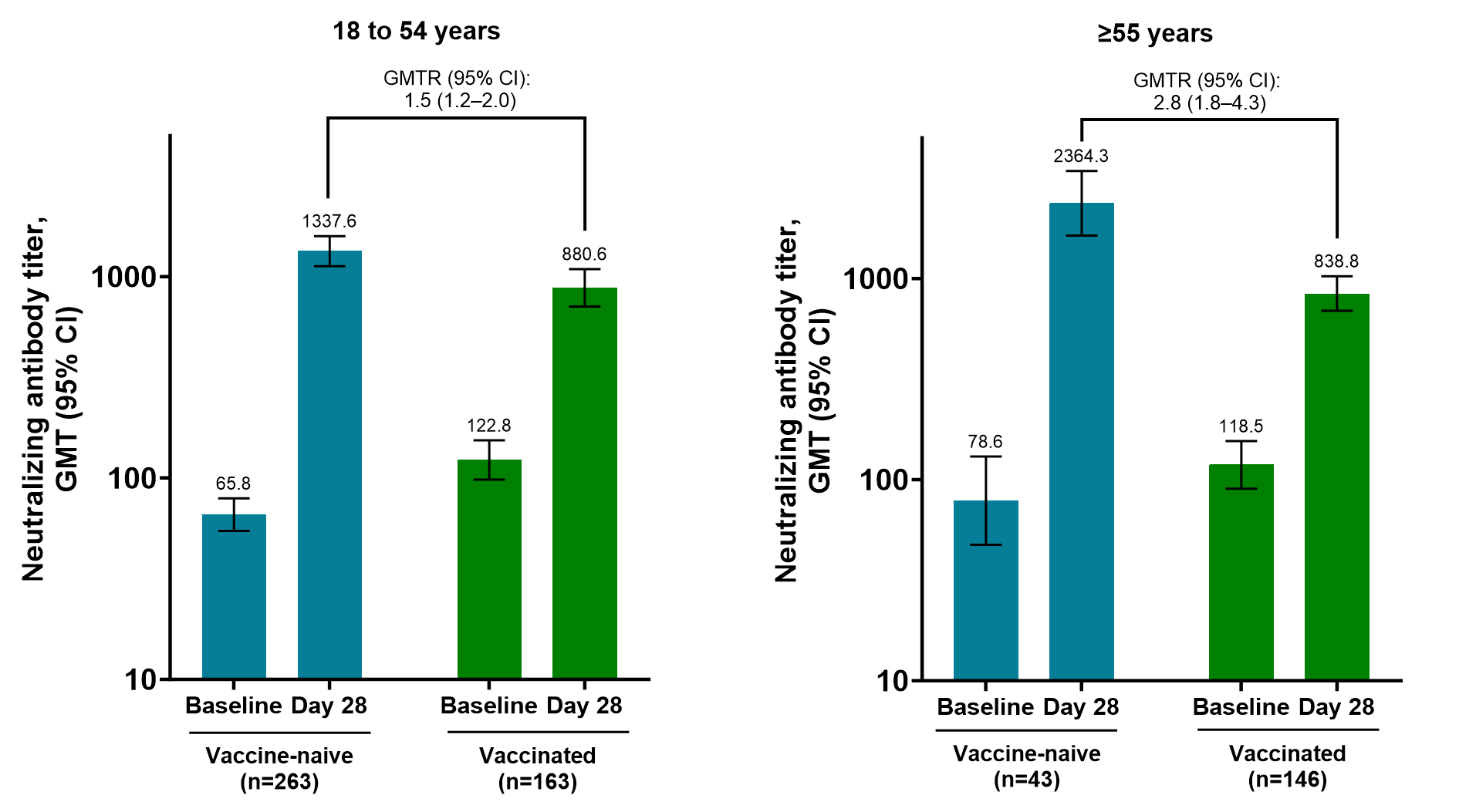

**Figure S2.** Neutralising antibody responses to XBB.1.5 at days 0 and 28 post study vaccination by age group (per-protocol analysis set)

GMTs of neutralising antibody responses to XBB.1.5 in vaccine naive and previously vaccinated participants. GMT values are shown on a log scale *y*-axis, and corresponding adjusted GMT values are shown above each respective bar. GMTRs (95% CI) are included comparing bracketed bars.

|  | **Vaccine-naive**  **(N=338)** | **Vaccinated**  **(N=332)** |
| --- | --- | --- |
| **Age, years** |  |  |
| Mean (SD) | 40.5 (13.11) | 52.0 (16.07) |
| Median (IQR) | 38 (31–49) | 53 (40–65) |
| **Age, category, n (%)** |  |  |
| 18 to 54 years | 284 (84.0) | 176 (53.0) |
| ≥55 years | 54 (16.0) | 156 (47.0) |
| **Sex, n (%)** |  |  |
| Female | 190 (56.2) | 208 (62.7) |
| Male | 148 (43.8) | 124 (37.3) |
| **Race, n (%)** |  |  |
| White | 167 (49.4) | 248 (74.7) |
| Black/African American | 147 (43.5) | 53 (16.0) |
| Asian | 2 (0.6) | 12 (3.6) |
| Native American/Alaska Native | 6 (1.8) | 6 (1.8) |
| Native Hawaiian/other Pacific Islander | 1 (0.3) | 2 (0.6) |
| Mixed origin | 5 (1.5) | 3 (0.9) |
| Other | 3 (0.9) | 1 (0.3) |
| Unknown/not reported | 7 (2.1) | 7 (2.1) |
| **Ethnicity, n (%)** |  |  |
| Not Hispanic/Latino | 249 (73.7) | 261 (78.6) |
| Hispanic/Latino | 87 (25.7) | 67 (20.2) |
| Not reported | 2 (0.6) | 4 (1.2) |
| Unknown | 0 | 0 |
| **Previous COVID-19, n (%)*** | 333 (98.5) | 5 (1.5) |
| **PCR, n (%)** |  |  |
| Negative | 333 (98.5)^†^ | 327 (98.5)^‡^ |
| Positive | 5 (1.5) | 5 (1.5) |
| **Anti-N/PCR status, n (%)^§^** |  |  |
| Positive | 315 (93.2) | 234 (70.5) |
| Negative | 23 (6.8) | 98 (29.5) |

**Table S1.** Participant baseline demographics and characteristics in the safety analysis set

*Previous COVID-19 occurrence was obtained via participant disclosure at enrollment.

^†^Five participants with missing PCR at baseline were imputed as negative in part 2.

^‡^Two participants with missing PCR at baseline were imputed as negative in part 1.

**^§^**Participants with a positive result for either anti-N or PCR are reported.

|  | **Vaccine-naive (N=306)** | **Vaccinated (N=309)** |
| --- | --- | --- |
| n | 288 | 305 |
| Baseline GMEU (95% CI)^†^ | 5218.1 (4339.6–6274.5) | 23701.1 (21042.3–26695.8) |
| Day-28 GMEU (95% CI) | 47045.1 (42090.2–52583.3) | 83205.7 (74747.0–92621.6) |
| Day-28 adjusted GMEU (95% CI) | 60691.2 (54585.2–67480.3) | 79761.5 (73603.5–86434.7) |
| GMFR from baseline to day 28 (95% CI) | 9.0 (7.6–10.7) | 3.5 (3.2–3.9) |
| Day-28 SRR, % (95% CI) | 68.4 (62.7–73.7) | 39.7 (34.1–45.4) |
| GMEUR (95% CI) | 0.9 (0.8–1.1) | |
| Difference in SRR, % (95% CI) | 28.7 (20.9–36.2) | |

**Table S2.** Anti-rS IgG responses* against XBB.1.5 (Per-protocol analysis set)

*The 95% CIs for GMEU and GMFR are calculated based on the t-distribution of the log-transformed values, then back transformed to the original scale.

^†^Baseline is the last non-missing result before study vaccination.

| **System organ class***  **Preferred term, n participants (%)** | **Vaccine-naive** | | | **Vaccinated** | | |
| --- | --- | --- | --- | --- | --- | --- |
|  | **Any** | **SAE** | **MAAE** | **Any** | **SAE** | **MAAE** |
| Any | 18 (5.3) | 1 (0.3) | 8 (2.4) | 29 (8.7) | 2 (0.6) | 14 (4.2) |
| Infections and infestations | 4 (1.2) | 0 | 3 (0.9) | 12 (3.6) | 1 (0.3) | 6 (1.8) |
| Urinary tract infection | 1 (0.3) | 0 | 1 (0.3) | 1 (0.3) | 0 | 0 |
| Cellulitis | 1 (0.3) | 0 | 1 (0.3) | 0 | 0 | 0 |
| Otitis media | 1 (0.3) | 0 | 1 (0.3) | 0 | 0 | 0 |
| Oral herpes | 1 (0.3) | 0 | 0 | 0 | 0 | 0 |
| COVID-19 | 0 | 0 | 0 | 5 (1.5) | 0 | 1 (0.3) |
| Tooth abscess | 0 | 0 | 0 | 2 (0.6) | 0 | 2 (0.6) |
| Appendiceal abscess | 0 | 0 | 0 | 1 (0.3) | 1 (0.3)^‡^ | 1 (0.3) |
| Gastroenteritis (*E. coli*) | 0 | 0 | 0 | 1 (0.3) | 0 | 1 (0.3) |
| Pharyngitis | 0 | 0 |  | 1 (0.3) | 0 | 0 |
| Viral respiratory tract infection | 0 | 0 | 0 | 1 (0.3) | 0 | 1 (0.3) |
| Tonsillitis | 0 | 0 | 0 | 1 (0.3) | 0 | 1 (0.3) |
| Respiratory, thoracic, and mediastinal disorders | 3 (0.9) | 0 | 1 (0.3) | 4 (1.2) | 0 | 1 (0.3 |
| Cough | 2 (0.6) | 0 | 1 (0.3) | 2 (0.6) | 0 | 1 (0.3) |
| Oropharyngeal pain | 1 (0.3) | 0 | 0 | 0 | 0 | 0 |
| Rhinorrhea | 1 (0.3) | 0 | 0 | 0 | 0 | 0 |
| Asthma | 0 | 0 | 0 | 1 (0.3)^†^ | 0 | 0 |
| Productive cough | 0 | 0 | 0 | 1 (0.3) | 0 | 0 |
| Injury, poisoning, and procedural complications | 3 (0.9) | 0 | 2 (0.6) | 2 (0.6) | 0 | 1 (0.3) |
| Fractured coccyx | 1 (0.3) | 0 | 1 (0.3) | 0 | 0 | 0 |
| Road traffic accident | 1 (0.3) | 0 | 1 (0.3) | 0 | 0 | 0 |
| Joint injury | 1 (0.3) | 0 | 0 | 0 | 0 | 0 |
| Tooth fracture | 0 | 0 | 0 | 1 (0.3) | 0 | 1 (0.3) |
| Contusion | 0 | 0 | 0 | 1 (0.3) | 0 | 0 |
| Gastrointestinal disorders | 2 (0.6) | 1 (0.3) | 2 (0.6) | 1 (0.3) | 0 | 0 |
| Obstructive pancreatitis | 1 (0.3) | 1 (0.3)^‡^ | 1 (0.3) | 0 | 0 | 0 |
| Abdominal pain | 1 (0.3) |  | 1 (0.3) | 0 | 0 | 0 |
| Diarrhea | 0 | 0 | 0 | 1 (0.3)^†^ | 0 | 0 |
| Immune system disorders | 2 (0.6) | 0 | 1 (0.3) | 0 | 0 | 0 |
| Hypersensitivity | 1 (0.3) | 0 | 1 (0.3) | 0 | 0 | 0 |
| Seasonal allergy | 1 (0.3) | 0 | 0 | 0 | 0 | 0 |
| Psychiatric disorders | 2 (0.6) | 0 | 1 (0.3) | 0 | 0 | 0 |
| Anxiety | 1 (0.3) | 0 | 1 (0.3) | 0 | 0 | 0 |
| Generalized anxiety disorder | 1 (0.3) | 0 | 0 | 0 | 0 | 0 |
| General disorders and administrative site conditions | 2 (0.6) | 0 | 0 | 2 (0.6)^†^ | 0 | 0 |
| Chills | 1 (0.3) | 0 | 0 | 0 | 0 | 0 |
| Injection site erythema | 1 (0.3) | 0 | 0 | 0 | 0 | 0 |
| Injection site pain | 1 (0.3) | 0 | 0 | 0 | 0 | 0 |
| Injection site pruritis | 1 (0.3) | 0 | 0 | 0 | 0 | 0 |
| Axillary pain | 0 | 0 | 0 | 1 (0.3)^†^ | 0 | 0 |
| Noncardiac chest pain | 0 | 0 | 0 | 1 (0.3) | 0 | 0 |
| Musculoskeletal and connective tissue disorders | 2 (0.6) | 0 | 0 | 0 | 0 | 0 |
| Jaw cyst | 1 (0.3) | 0 | 0 | 0 | 0 | 0 |
| Neck pain | 1 (0.3) | 0 | 0 | 0 | 0 | 0 |
| Pain in extremity | 1 (0.3) | 0 | 0 | 0 | 0 | 0 |
| Nervous system disorder | 1 (0.3) | 0 | 0 | 4 (1.2) | 0 | 2 (0.6) |
| Paresthesia | 1 (0.3) | 0 | 0 | 0 | 0 | 0 |
| Migraine | 0 | 0 | 0 | 2 (0.6) | 0 | 2 (0.6) |
| Presyncope | 0 | 0 | 0 | 1 (0.3)^†^ | 0 | 0 |
| Tension headache | 0 | 0 | 0 | 1 (0.3) | 0 | 0 |
| Reproductive system and breast disorders | 1 (0.3) | 0 | 0 | 0 | 0 | 0 |
| Heavy menstrual bleeding | 1 (0.3)^†^ | 0 | 0 | 0 | 0 | 0 |
| Investigations | 0 | 0 | 0 | 2 (0.6) | 0 | 1 (0.3) |
| Blood estrogen decreased | 0 | 0 | 0 | 1 (0.3) | 0 | 1 (0.3) |
| Body temperature increased | 0 | 0 | 0 | 1 (0.3) | 0 | 0 |
| Vascular disorders | 0 | 0 | 0 | 2 (0.6) | 0 | 2 (0.6) |
| Hypertension | 0 | 0 | 0 | 2 (0.6)^†^ | 0 | 2 (0.6) |
| Neoplasms | 0 | 0 | 0 | 1 (0.3) | 1 (0.3) | 1 (0.3) |
| Gastrointestinal stromal tumor | 0 | 0 | 0 | 1 (0.3) | 1 (0.3)^‡^ | 1 (0.3) |
| Blood and lymphatic disorders | 0 | 0 | 0 | 1 (0.3) | 0 | 0 |
| Anemia | 0 | 0 | 0 | 1 (0.3) | 0 | 0 |

**Table S3.** Unsolicited TEAEs through 28 days post study vaccination (Safety analysis set)

*MedDRA version 25.0. ^†^Event was considered treatment-related by the study investigator (heavy menstrual bleeding, diarrhea, axillary pain, presyncope, asthma, and one case of hypertension). ^‡^The events of obstructive pancreatitis, appendiceal abscess, and gastrointestinal stromal tumor were also reported as severe TEAEs.

| **First name** | **Last name** | **Affiliation** |
| --- | --- | --- |
| Jeffrey | Adelglass | (Elite) Research Your Health |
| Adebayo | Akinsola | Tekton Research |
| Brandon | Alleman | Tekton Research |
| Codey | Bell | Tekton Research |
| Laurence | Chu | Benchmark Research |
| Matthew | Davis | Rochester Clinical Research |
| Sue | Fanning | AMR |
| David | Ferrera | Benchmark Research |
| George | Freeman | Health Research of Hampton Roads, Inc |
| Linda | Gorgos | (Elite) AXCES Research |
| Ripley | Hollister | (Elite) Lynn Institute of the Rockies |
| Michael | Jacobs | AMR |
| Craig | Julien | AMR |
| Karen | Kotloff | University of Maryland |
| Robert | Lockwood | Tekton Research |
| R. Scott | McClelland | University of Washington |
| Jara | McDonald | Tekton Research |
| Abel | Murillo | AMR |
| Robert | Noveck | AMR |
| Paul | Pickrell | Tekton Research |
| William | Seger | Benchmark Research |
| Stacy | Slechta | AMR |
| William | Smith | AMR |
| Harry | Studdard | AMR |
| Ronald | Surowitz | (Elite) Health Awareness, Inc. |
| Milagritos | Tapia | University of Maryland |
| Eduardo | Uribe | PanAmerican Clinical Research |
| Keith | Vrbicky | Velocity Clinical Research |
| Larkin | Wadsworth | Sundance Clinical Research, LLC |
| Kem | Yenal | (Elite) DM Clinical Research - Philadelphia |
| Pedro | Ylisastigui | AMR |

**2019nCoV-313 study investigators**
